# Automated severe aortic stenosis detection on single-view echocardiography: A multi-center deep learning study

**DOI:** 10.1101/2022.08.30.22279413

**Authors:** Gregory Holste, Evangelos K. Oikonomou, Bobak J. Mortazavi, Andreas Coppi, Kamil F. Faridi, Edward J. Miller, John K. Forrest, Robert L. McNamara, Lucila Ohno-Machado, Neal Yuan, Aakriti Gupta, David Ouyang, Harlan M. Krumholz, Zhangyang Wang, Rohan Khera

## Abstract

**Background and Aims:** Early diagnosis of aortic stenosis (AS) is critical to prevent morbidity and mortality but requires skilled examination with Doppler imaging. This study reports the development and validation of a novel deep learning model that relies on 2-dimensional parasternal long axis (PLAX) videos from transthoracic echocardiography (TTE) without Doppler imaging to identify severe AS, suitable for point-of-care ultrasonography.

**Methods:** In a training set of 5,257 studies (17,570 videos) from 2016-2020 (Yale-New Haven Hospital [YNHH], Connecticut), an ensemble of 3-dimensional convolutional neural networks was developed to detect severe AS, leveraging self-supervised contrastive pretraining for label-efficient model development. This deep learning model was validated in a temporally distinct set of 2,040 consecutive studies from 2021 from YNHH as well as two geographically distinct cohorts of 5,572 and 865 studies, from California and other hospitals in New England, respectively.

**Results:** The deep learning model achieved an AUROC of 0.978 (95% CI: 0.966, 0.988) for detecting severe AS with 95.4% specificity and 90% sensitivity in the temporally distinct test set, maintaining its diagnostic performance in both geographically distinct cohorts (AUROC 0.972 [95% CI: 0.969, 0.975] in California and 0.915 [95% CI: 0.896, 0.933] in New England, respectively). The model was interpretable with saliency maps identifying the aortic valve as the predictive region. Among non-severe AS cases, predicted probabilities were associated with worse quantitative metrics of AS suggesting association with various stages of AS severity.

**Conclusions:** This study developed and externally validated an automated approach for severe AS detection using single-view 2D echocardiography, with implications for point-of-care screening.

**STRUCTURED GRAPHICAL ABSTRACT:** *Key Question:* Is it feasible to automatically screen for the presence of severe aortic stenosis (AS) using single-view echocardiographic videos without the use of Doppler imaging?

*Key Finding:* Using self-supervised pretraining and ensemble learning, we trained a deep learning model to detect severe AS using single-view echocardiography without Doppler imaging. The model maintained its high performance in multiple geographically and temporally distinct cohorts.

*Take-home Message:* We present an automated method to detect severe AS using single-view TTE videos, with implications for point-of-care ultrasound screening as part of routine clinic visits and in limited resource settings by individuals with minimal training. 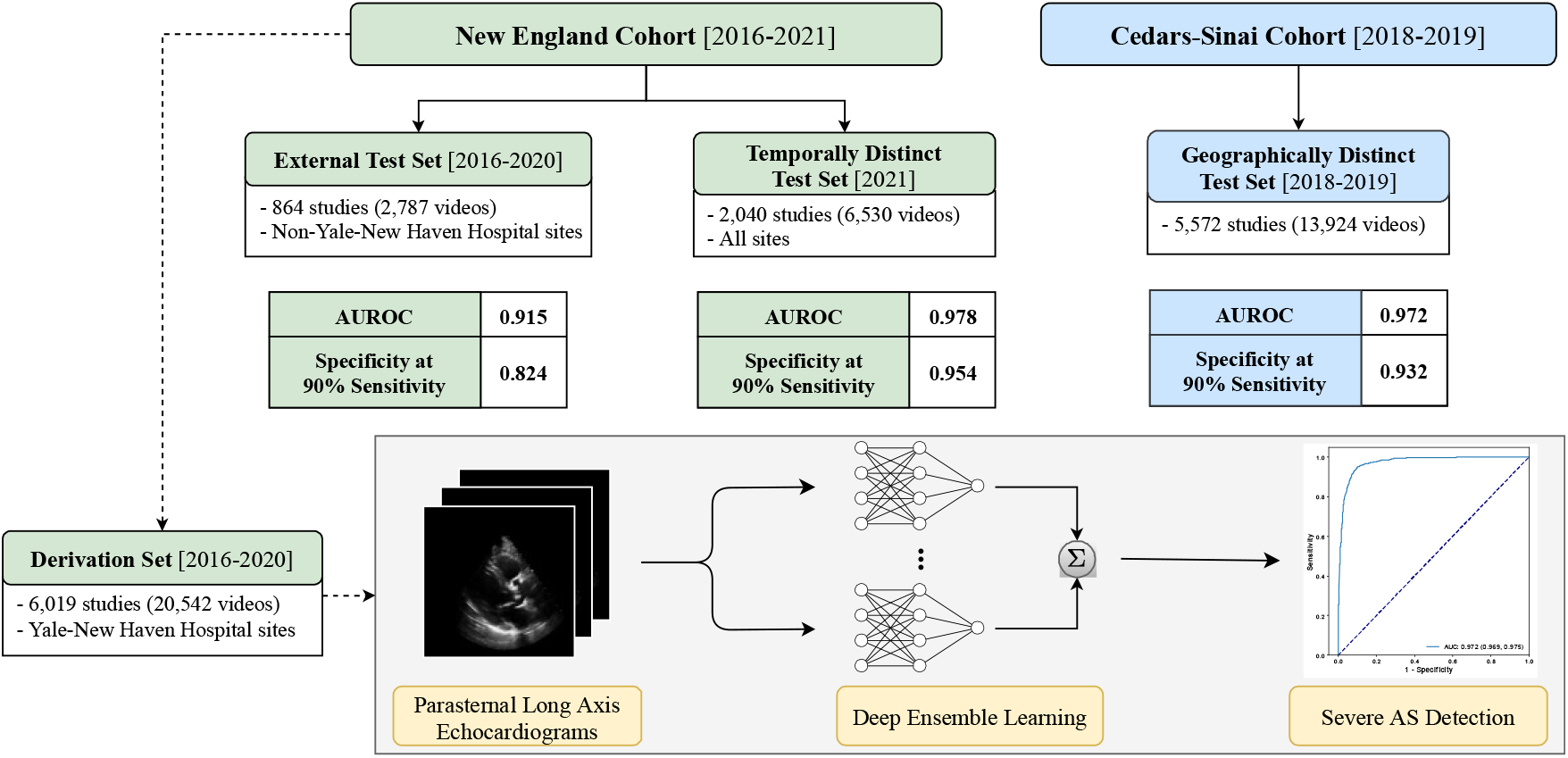 An automated deep learning approach for severe aortic stenosis detection from single-view echocardiography evaluated across geographically and temporally distinct cohorts.

## INTRODUCTION

Aortic stenosis (AS) is a chronic, progressive disease, and associated with morbidity and mortality.^1,2^ With advances in both surgical and transcatheter aortic valve replacement,^3^ there has been an increasing focus on early detection and management.^4,5,6^ The non-invasive diagnosis of AS can be made with hemodynamic measurements using Doppler echocardiography,^2,7,8^ but that requires dedicated equipment and skilled acquisition and interpretation. On the other hand, even though two-dimensional (2D) cardiac ultrasonography is increasingly available with handheld devices that can visualize the heart,^9^ it has not been validated for the diagnosis or longitudinal monitoring of AS. With an estimated prevalence of 5% among individuals aged 65 years or older,^8^ there is a growing need for user-friendly screening tools which can be used in everyday practice by people with minimal training to screen for severe AS. This need for timely screening is further supported by evidence suggesting improved outcomes with early intervention even in the absence of symptomatic disease.^5^

Machine learning offers opportunities to standardize the acquisition and interpretation of medical images.^10^ Deep learning algorithms have successfully been applied in echocardiograms, where they have shown promise in detecting left ventricular dysfunction,^11^ and left ventricular hypertrophy.^12^ With the expanded use of point-of-care ultrasonography,^9^ developing user-friendly screening algorithms relying on single 2D echocardiographic views would provide an opportunity to improve AS screening by operators with minimal experience through time-efficient protocols. This is often limited by the lack of carefully curated, labelled datasets, as well as efficient ways to utilize the often noisy real-world data for model development.^13^

In the present study, we hypothesized that a deep learning model trained on 2D echocardiographic views of parasternal long axis (PLAX) videos can reliably predict the presence of severe AS without requiring Doppler input. The approach leverages self-supervised learning of PLAX videos along with two other neural network initialization methods to form a diverse ensemble model capable of identifying severe AS from raw 2D echocardiograms. The model is trained based on a dataset from different operators and machines, with its external performance assessed both in geographically and temporally distinct cohorts. Combined with automated view classification, our approach serves as an end-to-end automated solution for deep learning applications in the field of point-of-care echocardiography.

## METHODS

### Study population & data source

#### New England cohort (Yale-New Haven Health network)

A total of 12,500 studies were queried from all TTE exams performed between 2016 and 2021 across the Yale New Haven Health System (YNHHS, including Connecticut and Rhode Island), and were used for model derivation & testing across different hospitals and time periods. For internal model development and evaluation, 10,000 studies from 2016-2020 were randomly queried with AS oversampled to mitigate class imbalance during model training. Specifically, this query sampled normal studies uniformly (including “no AS” and “sclerosis without stenosis”), oversampled non-severe AS studies by 5-fold (including “mild AS”, “mild-moderate AS”, “moderate-severe AS”, “low gradient AS”, and “paradoxical AS”), and oversampled severe AS by 50-fold. This strategy was designed to ensure that the model encounters sufficient examples of severe AS to learn the signatures of the disorder. The 10,000 studies were then split at the patient level into a derivation set (consisting of all patients scanned in the Yale-New Haven Hospital, Connecticut, USA, including satellite locations) and a geographically distinct, external testing set from New England (consisting of patients scanned at four other hospitals – namely Bridgeport Hospital, Lawrence & Memorial Hospital, and Greenwich Hospital, all in Connecticut, USA – as well as the Westerly hospital in Rhode Island, USA). The remaining 2,500 studies of the query were all conducted across the previously mentioned centers a year later in 2021 with *no oversampling* to serve as a challenging temporally distinct testing set, where severe AS represents approximately 1% of all cases. The study population is summarized in **Figure 1**.

**Figure 1.**
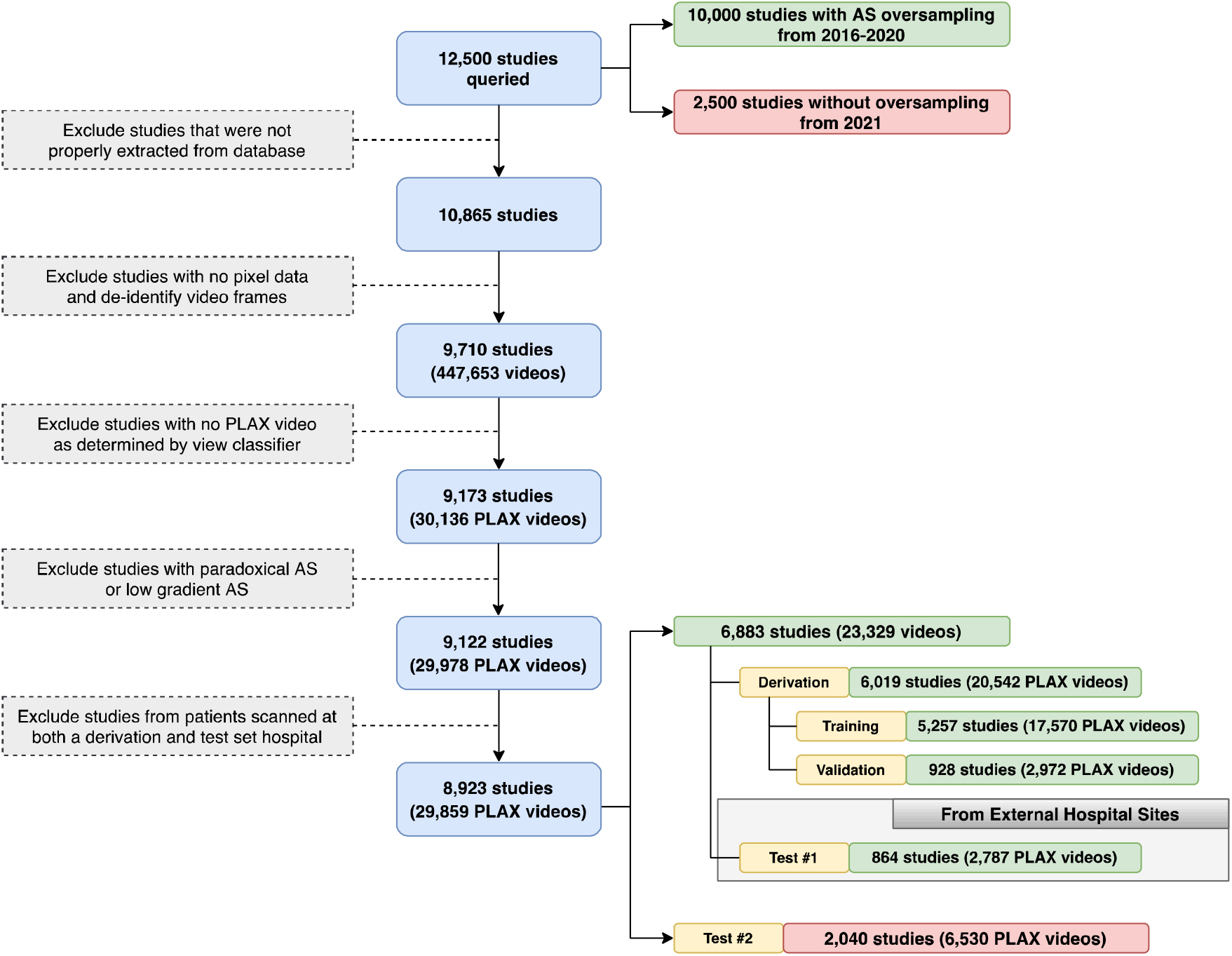
Inclusion-exclusion flowchart for the New England study population. Exclusion criteria for transthoracic echocardiogram (TTE) studies and videos included in this study from the Yale-New Haven Health network. Studies with valid pixel data were de-identified frame by frame, and the parasternal long axis (PLAX) view was determined by an automated view classifier. A sample of 10,000 studies from 2016-2020 (with AS oversampled) were split into a derivation set and external test set, which comprised studies from hospital sites not encountered during model training. An independent random sample of 2,500 studies from 2021 (with no oversampling) was used as an additional test set to evaluate robustness to temporal shift.

#### Cedars-Sinai cohort

For further testing in an additional geographically distinct cohort, all transthoracic echocardiograms performed at the Cedars-Sinai Medical Center (Los Angeles, California, USA) between January 1^st^ 2018 and December 31^st^ 2019 were retrieved. AS severity was determined from finalized TTE reports. After excluding studies with prosthetic aortic valves, 3,867 TTEs without severe aortic stenosis were sampled at random and combined with 1,705 TTEs with severe AS to create an enriched 5,572-study cohort.

#### Consent

The study was reviewed by the Yale and Cedars-Sinai Institutional Review Boards, which approved the study protocol and waived the need for informed consent as the study represents secondary analysis of existing data.

#### Echocardiogram interpretation

All studies were performed by trained echocardiographers or cardiologists and reported by board-certified cardiologists with specific training cardiac echocardiography. These reports were a part of routine clinical care, in accordance with the recommendations of the American Society of Echocardiography (ASE).^14,15^ The presence of AS severity was adjudicated based on the original echocardiographic report. Further details on the measurements obtained are presented in the ***Supplement***.

### Model training & development

#### Data pre-processing

All studies underwent de-identification, view classification, and preprocessing to curate a dataset of PLAX videos for deep-learned severe AS prediction. The full process describing the extraction of the echocardiographic videos, loading of image data, masking of identifying information, conversion to Audio Video Interleave format (AVI), and downsampling for further processing and automated view classification are described in the ***Supplement***. Briefly, after excluding studies that were not properly extracted or contained no pixel data, 9,710 studies with 447,653 videos underwent automated view classification based on a pretrained TTE view classifier.^16^ We retained videos where the automated view classifier most confidently predicted the presence of a PLAX view. After excluding cases of low-flow, low-gradient, and paradoxical AS (determined based on the final clinical report), the final Yale-New Haven Health system dataset consisted of 30,136 videos in 9,173 studies. This included 2,040 studies from 2021, which were set aside to form a temporally distinct testing set, while the remaining 7,082 studies (with AS oversampled as described above) were split into a derivation set (Yale-New Haven Hospital and satellite centers) and a geographically distinct testing set (see *Study population & data source*, **Figure 1**). To ensure no patient overlap between derivation and testing sets, 199 studies corresponding to patients in the derivation set were not included in the testing set. Finally, the derivation set was randomly split into training and validation sets at an 85%/15% ratio, leaving 5,257 studies for training, 928 studies for internal validation during training, and 864 studies for external validation.

#### Self-supervised learning (SSL)

We used our previously described novel approach of self-supervised contrastive pretraining for echocardiogram videos.^17^ This approach demonstrated that classification tasks could be performed in a more data-efficient manner through “in-domain” pretraining on echocardiograms,^17^ as opposed to other standard approaches such as random initialization of weights and transfer learning.^11,18,19^ Briefly, this self-supervised learning was performed on the training set videos with a novel combination of (i) a multi-instance contrastive learning task and (ii) a frame re-ordering pretext task, both explained in detail in the ***Supplement*** and summarized in **Figure 2**. For this, we adopted “multi-instance” contrastive learning, where the model was trained to learn similar representations of *different* videos from the *same* patient, which allowed the model to learn the latent space of PLAX-view echocardiographic videos. To additionally encourage temporal coherence of our model, we included a frame re-ordering “pretext” task to our self-supervised learning method, where we randomly permuted the frames of each input echo, then trained the model to predict the original order of frames.^22^

**Figure 2.**
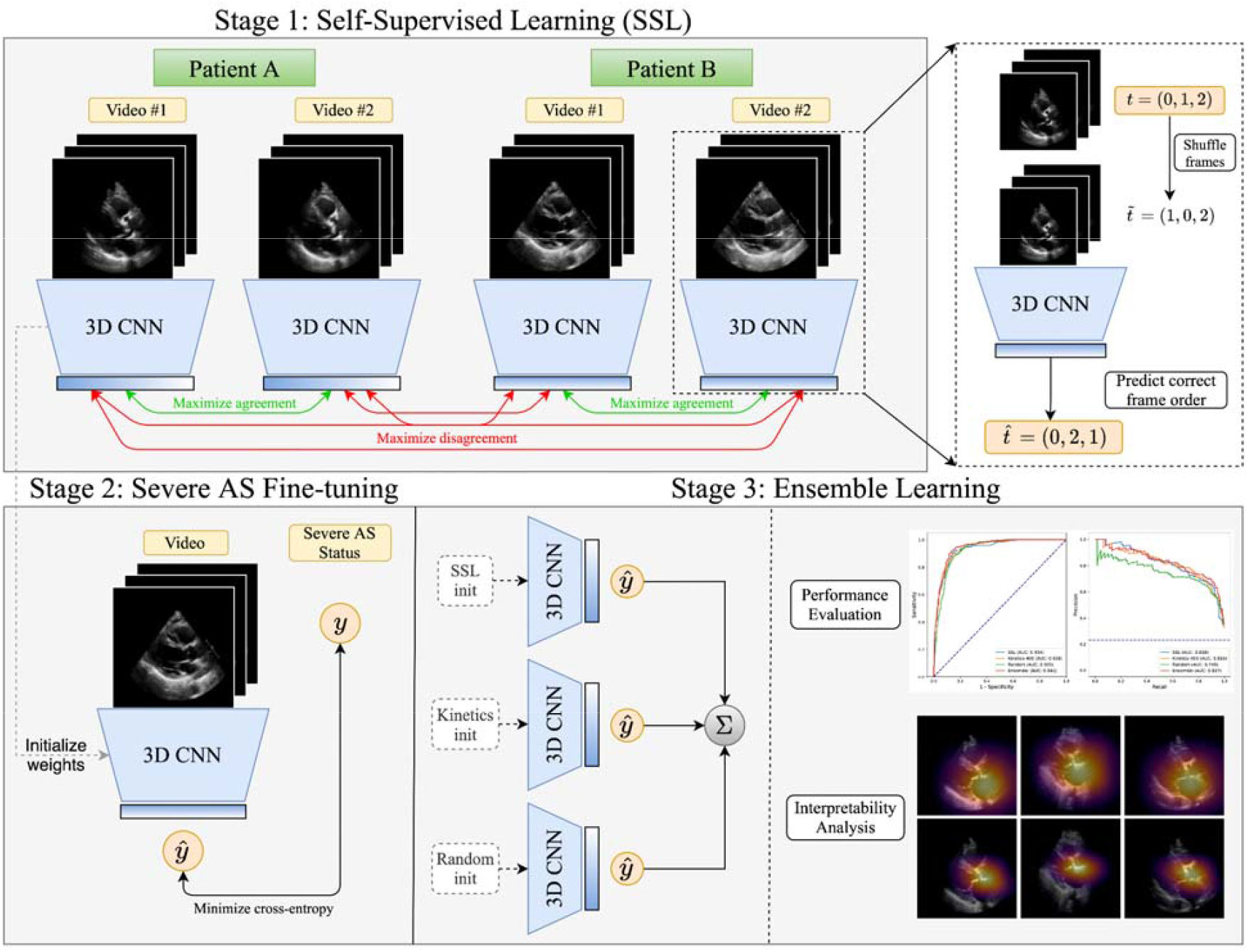
Overview of proposed approach. We first perform self-supervised pretraining on parasternal long axis (PLAX) echocardiogram videos, selecting different PLAX videos from the same patient as “positive samples” for contrastive learning. After this representation learning step, we then use these learned weights as the initialization for a model that is fine-tuned to predict severe aortic stenosis (AS) in a supervised fashion.

Self-supervised pretraining was performed on randomly sampled video clips of 4 consecutive frames from each of the training set echocardiogram videos for 300 epochs. A 3D-ResNet18^23^ architecture was used as the encoder (see the ***Supplement*** for full architecture details). The model was trained for 300 epochs on all unique pairs of different PLAX videos from the same patient with the Adam optimizer,^24^ a learning rate of 0.1, a batch size of 392 (196 per GPU), and NT-Xent temperature hyperparameter of 0.5. The following augmentations were applied to each frame in a temporally consistent manner (same transformations for each frame of a given video clip): random zero padding by up to 8 pixels in each spatial dimension, a random horizontal flip with probability 0.5, and a random rotation within -10 and 10 degrees with probability 0.5.

#### Deep neural network training for severe AS prediction

The 3D-ResNet18 architecture described above was also leveraged to detect severe AS. Three different methods were used to initialize the parameters of this network: an SSL initialization, a Kinetics-400 initialization, and a random initialization (see ***Supplement***, and **Figure 2**). All fine-tuning models were trained on randomly sampled video clips of 16 consecutive frames from training set echocardiograms.

Models were trained for a maximum of 30 epochs with early stopping if validation AUROC did not improve for 5 consecutive epochs. Severe AS models were trained on a single NVIDIA RTX 3090 GPU with the Adam optimizer, a learning rate of 1 × 10^−4^ (except the SSL-pretrained model, which used a learning rate of 0.1) and a batch size of 88 using a sigmoid cross-entropy loss. We additionally used class weights computed with the method provided by *scikit-learn*^25^ to accommodate class imbalance in addition to label smoothing^26,27^ with a=0.1, a method to improve model calibration and generalization. Learning curves depicting loss throughout training can be found in **Figure S1**.

#### Ensemble learning

Since models were trained on 16-frame video clips, we averaged clip-level predictions to obtain video-level predictions of severe AS. After repeating the process for all videos, severe AS probabilities for all videos were then averaged to obtain study-level AS predictions. The final ensemble model is then formed by averaging the study-level output probabilities of the SSL-pretrained model, the Kinetics-400-pretrained model, and the randomly initialized model after fine-tuning each ensemble member to detect severe AS. Since no quality control is applied when selecting PLAX videos for this work, averaging results over multiple videos in the same study has a stabilizing effect that boosts predictive performance.^28^

#### Assessing diagnostic performance in the testing sets

We evaluate the model’s performance on both area under the receiver operating characteristic curve (AUROC) and the area under the precision-recall curve (AUPRC), with the latter being specifically informative when class imbalance is present.^29^ We additionally reported metrics that assess performance at specific decision thresholds such as F1 score, positive predictive value (PPV), specificity at 90% sensitivity, and PPV at 90% sensitivity. For these metrics, we proceed with the threshold that maximizes F1 score, the harmonic mean of precision and recall, on the given evaluation set. The latter two metrics – specificity and PPV at 90% sensitivity – were included to provide a clinically relevant assessment of the model’s performance at the minimum sensitivity required for real-world deployment.

#### Model explainability

We evaluated the predictive focus of the models using saliency maps. These were generated using the Grad-CAM method^30^ for obtaining visual explanations from deep neural networks (see ***Supplement***). This method was used to produce a frame-by-frame “visual explanation” of where the model is focusing to make its prediction. To generate a single 2-dimensional heatmap for a given echo clip, the pixelwise maximum along the temporal axis was taken to capture the most salient regions for severe AS predictions across all timepoints.

These spatial attention maps were visualized based on the randomly initialized, Kinetics-400-pretrained, and SSL-pretrained AS models for five true positives, a true negative, and a false positive.

## Statistical analysis

All 95% confidence intervals for model performance metrics were computed by bootstrapping. Specifically, 10,000 bootstrap samples (samples with replacement having equal sample size to the original evaluation set) of the evaluation set were drawn, metrics were computed on this set of studies, and nonparametric confidence intervals were constructed with the percentile method. Bootstrapping was performed at the study level since the severe AS labels are provided for each echocardiographic study. For analysis of correlation between model outputs and quantitative measures of AS, categorical variables were summarized as percentages, whereas continuous variables are reported as mean values with standard deviation and visualized using violin plots. Continuous variables between two groups were compared using the Student’s *t*-test. Pearson’s *r* was used to assess the pairwise correlation between continuous variables. Spearman’s rank-order correlation test was used to analyze the relationship between model outputs and AS severity, which was represented ordinally (0=none, 1=mild-moderate, 2=severe). All statistical tests were two-sided with a significance level of 0.05, unless specified otherwise. Analyses were performed using Python (version 3.8.5).

## RESULTS

### Study Population

In the New England cohort, after removing studies with no pixel data, de-identifying video frames, and using an automated view classifier to determine the PLAX view, our final derivation set consisted of 6,019 studies with 20,542 videos (1,294,197 frames) (mean age 69.6 ± 15.7 years, n=2,992 [48.4%] women), with mild, moderate, and severe AS in 13.8% (n=780), 8.8% (n=495), and 19.2% (n=1,588) of studies, respectively. To evaluate generalization across local hospital sites, we curated a test set of 864 studies (2,787 videos) from separate New England hospitals in the YNHHS network that were not present in the derivation set. The temporally distinct test set consisted of 2,040 randomly selected scans with a total of 6,530 videos performed between January 1^st^ 2021 and December 15^th^ 2021 across YNHHS (mean age 65.7 ± 16.4 years, n=997 (48.9%) women) were used for time-dependent model validation. This temporally distinct set was *not* oversampled, with mild, moderate, and severe AS estimated in 4.1% (n=83), 2.9% (n=59), and 1.0% (n=20) of the studies, respectively. Finally, a set of 5,572 studies performed at the Cedars-Sinai Medical Center between 2018 and 2019, with oversampling for severe AS (1,705 studies with severe AS out of 5,572, 30.6%) (mean age 68.5 ± 17.1 years, n=2,370 [42.5%] women), was also used for further external testing (**Figure 1**). Further information on patient characteristics is presented in the **Methods** and **Table 1**.

**Table 1.** Table of baseline demographic and echocardiographic characteristics.

|  |  | New England (Yale-New Haven Health System) |  |  |  |  | California |
| --- | --- | --- | --- | --- | --- | --- | --- |
|  |  | Missing | Overall | Derivation | Geographically distinct testing #1 | Temporally distinct testing | Geographically distinct testing #2 |
| **n** |  |  | 9089 | 6185 | 864 | 2040 | 5572 |
| **Location** |  |  | - | YNHH | BH, GH, LMH, WH | YNHH, BH, GH, LMH, WH | CSMC |
| **Year of study** |  |  | - | 2016-2020 | 2016-2020 | 2021 | 2018-2019 |
| **Age (years), mean (SD)** |  | 6 | 69.1 (16.0) | 69.9 (15.7) | 70.9 (16.3) | 65.7 (16.4) | 68.5 (17.1) |
| **Gender, n (%)** | Female | 0 | 4453 (49.0) | 2992 (48.4) | 464 (53.7) | 997 (48.9) | 2370 (42.5) |
|  | Male |  | 4636 (51.0) | 3193 (51.6) | 400 (46.3) | 1043 (51.1) | 3202 (57.5) |
| **Race & Ethnicity (per echo reports), n (%)** | Asian | 6770 | 32 (1.4) | 21 (1.2) | 2 (1.6) | 9 (2.0) | 404 (7.3) |
|  | Black |  | 268 (11.6) | 188 (10.8) | 16 (12.9) | 64 (14.2) | 651 (11.8) |
|  | Hispanic |  | 118 (5.1) | 84 (4.8) | 10 (8.1) | 24 (5.3) | 556 (10.0) |
|  | Other |  | 37 (1.6) | 27 (1.5) | 4 (3.2) | 6 (1.3) | 451 (8.1) |
|  | Unknown |  | 18 (0.8) | 12 (0.7) | 2 (1.6) | 4 (0.9) | 101 (1.8) |
|  | White |  | 1846 (79.6) | 1411 (81.0) | 90 (72.6) | 345 (76.3) | 3409 (61.6) |
| **LVIDd Index (cm/m<sup>2</sup>), mean (SD)** |  | 1111 | 2.4 (0.4) | 2.4 (0.4) | 2.4 (0.4) | 2.4 (0.4) | 2.4 (0.6) |
| **RVSP (mmHg), mean (SD)** |  | 2500 | 32.3 (13.5) | 32.5 (13.3) | 35.7 (16.5) | 29.8 (12.0) | 32.8 (14.4) |
| **EF (%), mean (SD)** |  | 108 | 59.4 (10.8) | 59.5 (10.8) | 58.5 (12.0) | 59.1 (10.2) | 57.8 (14.8) |
| **AVA by VTI (cm<sup>2</sup>), mean (SD)** |  | 4713 | 1.4 (0.8) | 1.3 (0.8) | 1.5 (0.9) | 2.1 (0.9) | 1.4 (1.2) |
| **AV mean gradient (mmHg), mean (SD)** |  | 3649 | 20.5 (17.8) | 23.2 (18.2) | 18.4 (17.2) | 9.0 (9.4) | 23.8 (19.8) |
| **AV peak velocity (m/s), mean (SD)** |  | 441 | 2.2 (1.2) | 2.4 (1.3) | 2.2 (1.2) | 1.6 (0.6) | 2.3 (1.4) |

AV: aortic valve; BH: Bridgeport hospital; BP: blood pressure; CSMC: Cedars-Sinai Medical Center; EF: ejection fraction; GH: Greenwich hospital; LAD: left atrium; LMH: Lawrence & Memorial hospital; LVIDd: left ventricular internal diastolic diameter; RVSP: right ventricular systolic pressure; SD: standard deviation; VTI: velocity time integral, WH: Westerly hospital; YNHH: Yale-New Haven Hospital.

### Performance of a deep learning model for severe AS detection based on PLAX videos

The ensemble model was able to reliably detect the presence of severe AS using single-view, two-dimensional PLAX videos, demonstrating an AUROC of 0.915 (95% CI: 0.896, 0.933) and 82.4% sensitivity at 90% specificity (95% CI: 72.5%, 85.7%) on the geographically distinct testing set of New England hospitals not included in the derivation set. The model also demonstrated consistent performance across time in the same hospital system, maintaining its discriminatory performance with an AUROC of 0.978 (95% CI: 0.966, 0.988) and 95.4% sensitivity at 90% specificity (95% CI: 88.0%, 97.1%) on the temporally distinct testing set from 2021. Finally, in further geographically distinct testing using scans performed at Cedars-Sinai, the model generalized well across institutions, reaching 0.972 AUROC (95% CI: 0.969, 0.975) and 93.2% specificity at 90% sensitivity (95% CI: 92.4%, 94.0%). Since the prevalence of severe AS varied among the external testing populations, the AUPR varied from 0.414 (95% CI: 0.231, 0.594) in the temporally distinct test set (New England, 2021) to 0.932 (95% CI: 0.921, 0.941) in the external testing set from Cedars-Sinai. Receiver operating characteristic (ROC) and precision-recall (PR) curves, as well as the distribution of model probabilities across disease groups (no AS, mild-moderate AS, severe AS) showing a graded relationship across severity groups, are shown in **Figure 3**; see ***Supplemental Table S1*** for full detailed results. Furthermore, in sensitivity analyses without averaging predictions from multiple videos in the same study we observed overall consistent results, as summarized in ***Supplemental Table S2***.

**Figure 3.**
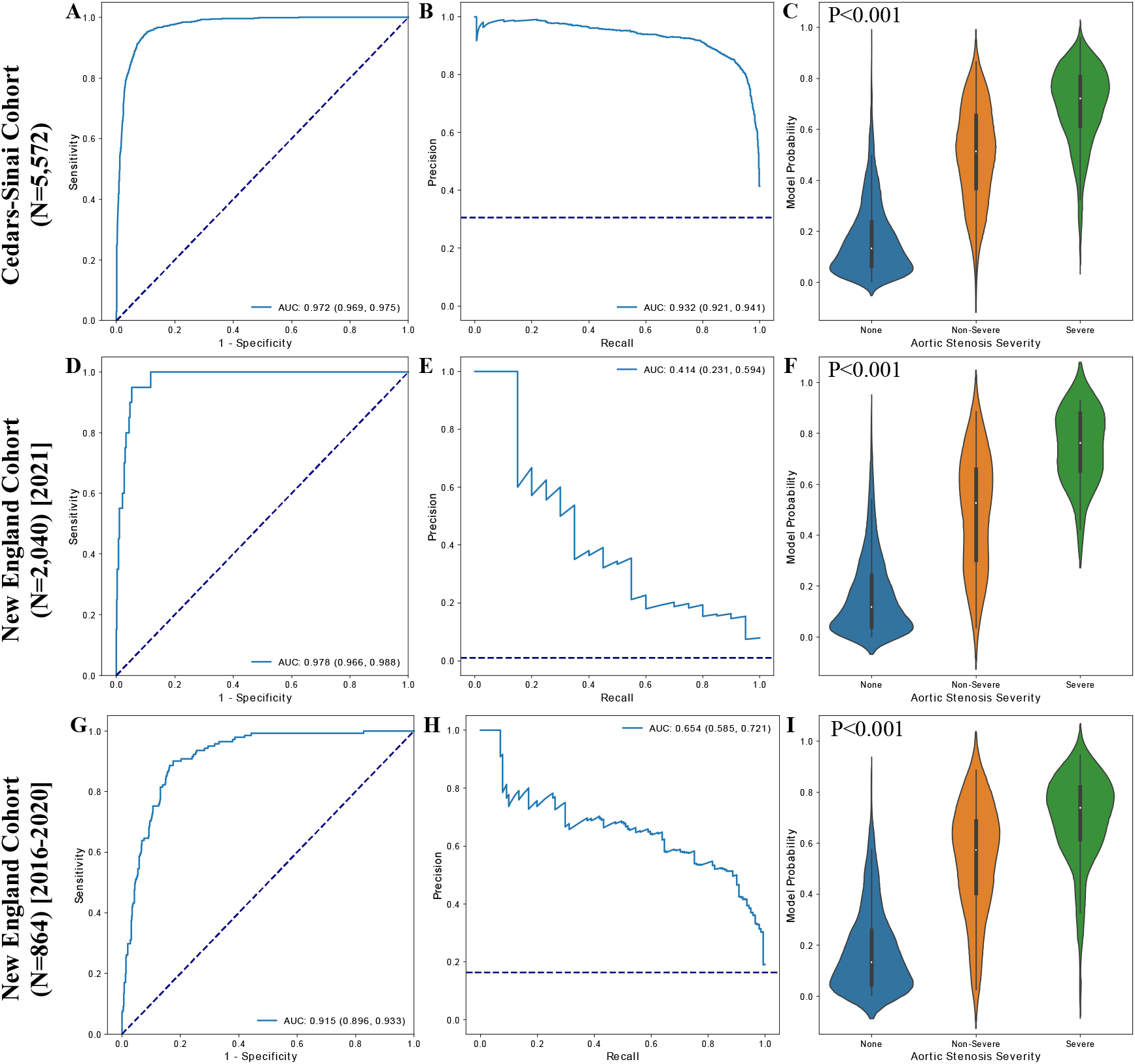
Model performance in the external validation sets. Receiver operating characteristic curves (first column), precision-recall curves (second column), and violin plots showing relationship of model output with aortic stenosis severity (third column) for the external Cedars-Sinai cohort (first row), temporally distinct New England cohort (second row), and external New England cohort (third row).

### Explainable predictions through saliency maps

We used Gradient-weighted Class Activation Mapping (Grad-CAM) to identify the regions in each video frame that contributed the most to the predicted label. In the examples shown in **Figure 4**, the first five columns represent the five most confident severe AS predictions, the sixth column represents the most confident “normal” (no severe AS) prediction, and the seventh column represents the most confident incorrect severe AS prediction. The saliency maps from our SSL approach demonstrated overall consistent and specific localization of the activation signal in the pixels corresponding to the aortic valve and annulus (bottom row).

**Figure 4.**
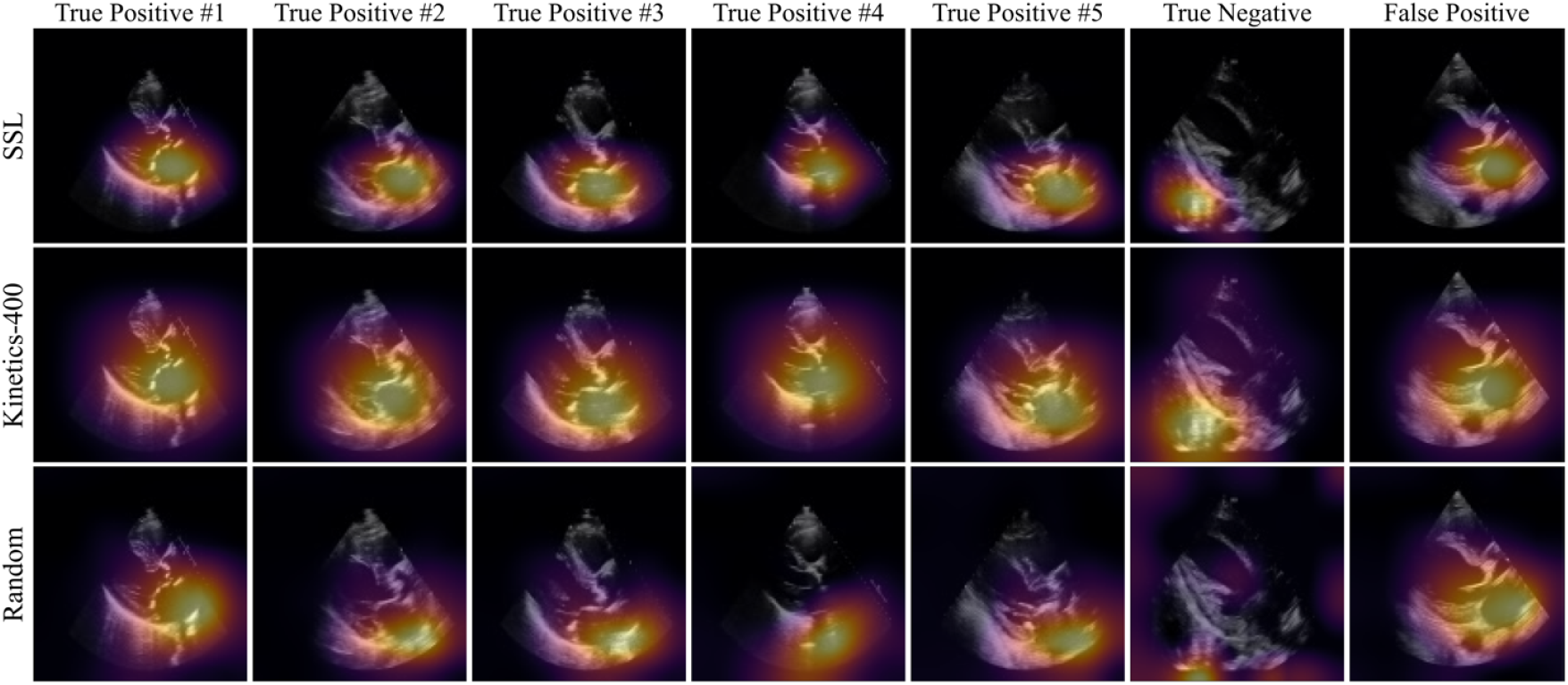
Saliency map visualization. Spatial attention maps for the randomly initialized model (top row), Kinetics-pretrained model (middle row) and self-supervised learning (SSL) approach (bottom row) for five true positives (first five columns), a true negative (sixth column), and a false positive (last column). As determined by the Kinetics-pretrained model, the first five columns represent the five most confident severe AS predictions, the sixth column represents the most confident “normal” (no severe AS) prediction, and the seventh column represents the most confident *incorrect* severe AS prediction. Saliency maps were computed with the GradCAM method and reduced to a single 2D heatmap by maximum intensity projection along the temporal axis.

### Model identification of features of AS severity

In the temporally distinct testing set from 2021 (reflecting the normal prevalence of severe AS in an echocardiographic cohort), we observed that the predictions of the ensemble model correlated with continuous metrics of AS severity, including the peak aortic valve velocity (r=0.59, P<0.001), trans-valvular mean gradient (r=0.66, P<0.001) and the mean aortic valve area (r=-0.53, P<0.001). On the other hand, the model predictions were independent of the left ventricular ejection fraction (LVEF) (r=-0.02, P=0.37), a negative control. In further sensitivity analysis, we stratified cases without AS or mild/moderate AS based on the predictions of our model as true negatives (TN) or false positives (FP). Compared to true negatives, false positive cases had significantly higher peak aortic velocities (FP: 3.4 [25^th^-75^th^ percentile: 2.9-3.7] m/sec; TN: 1.6 [1.3-2.3] m/sec, P<0.001), trans-valvular mean gradients (FP: 26.0 [25^th^-75^th^ percentile: 20.5-31.8] mmHg; TN: 5.0 [3.8-9.0] m/sec, P<0.001), and mean aortic valve area (FP: 1.04 [25^th^-75^th^ percentile: 0.86-1.28] cm^2^; TN: 1.99 [1.49-2.67] cm^2^, P<0.001), but no significant difference in the LVEF (FP: 65.4% [55.0%-67.8%]; TN: 60.0% [55.0-65.0%], P=0.19) (**Figure 5**).

**Figure 5.**
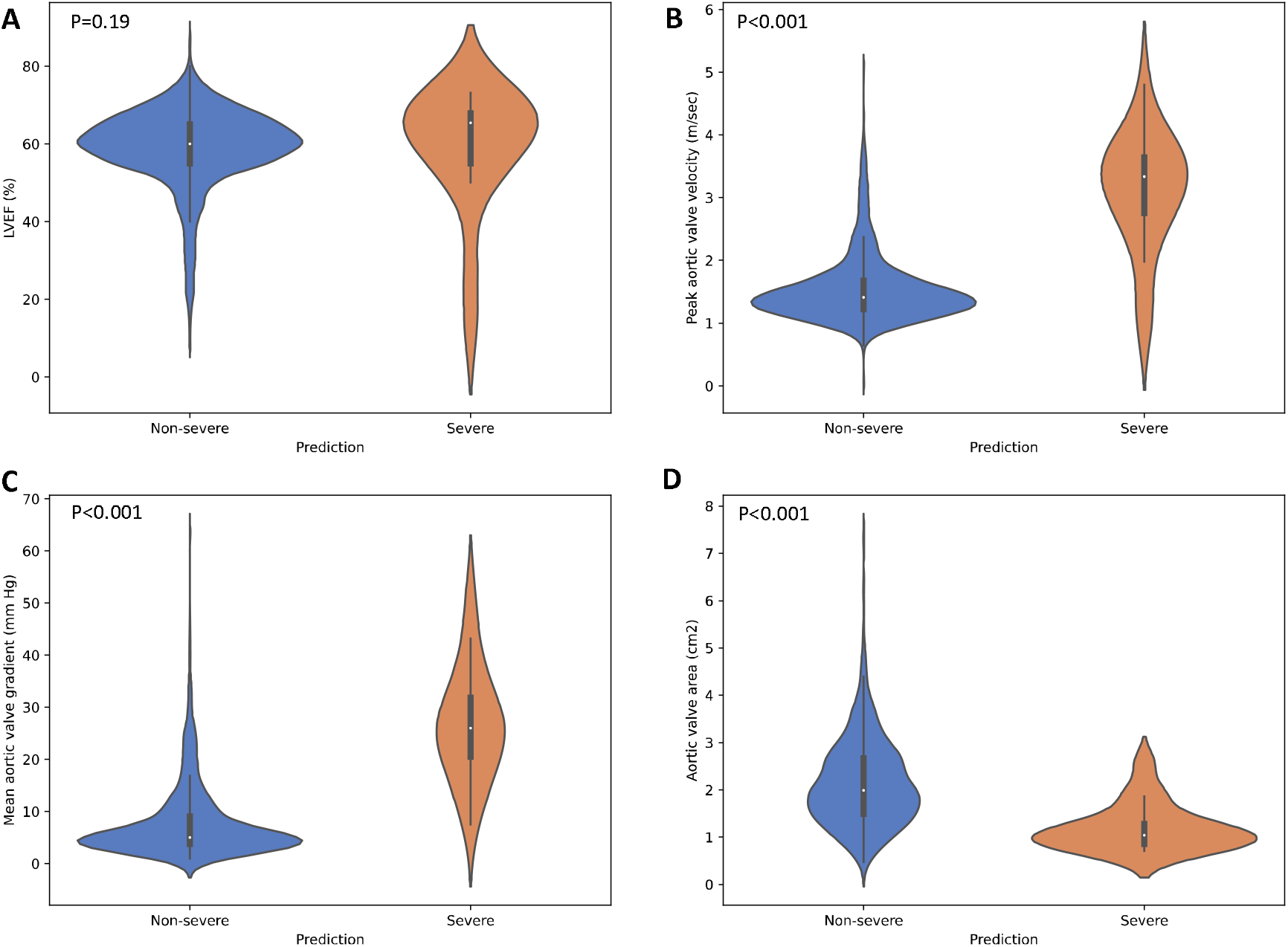
Comparison between model predictions and echocardiographic left ventricular and aortic valve assessment among patients without severe aortic stenosis. Violin plots demonstrating the distribution of LVEF (left ventricular ejection fraction, **A**) peak aortic valve velocity (**B**), mean aortic valve gradient (**C**) and mean aortic valve area (**D**) for patients without severe AS, stratified based on the predicted class based on the final ensemble model. These results are based on the temporally distinct cohort of patients scanned in 2021, without oversampling for severe aortic stenosis cases.

## DISCUSSION

We have developed and validated an automated algorithm that can efficiently screen for and detect the presence of severe AS based on a single-view two-dimensional transthoracic echocardiographic video. The algorithm demonstrates excellent performance (AUROC of 0.91 to 0.97), with high sensitivity (>93%) at high specificity (90%), maintaining its robustness and discriminatory performance across several geographically and temporally distinct cohorts with varying prevalence of severe AS. We also present a novel self-supervised step leveraging multi-instance contrastive learning, which allowed our algorithm to learn key representations that define each patient’s unique phenotype through contrastive pre-training, independent of the expected technical variation in image acquisition, including differences in probe orientation, beam angulation and depth. Visualization of saliency maps introduces explainability to our algorithms and confirms the key areas of the PLAX view, including the aortic valve and annulus, that contributed the most to our predictions. Furthermore, features learned by the model generalize to lower severity cases, highlighting the potential value of our model in the longitudinal monitoring of AS, a disease with a well-defined, progressive course.^2^ Our approach has the potential to expand the use of echocardiographic screening for suspected AS, shifting the burden away from dedicated echocardiographic laboratories to point-of-care screening in primary care offices, or low-resource settings. It may also enable operators with minimal echocardiographic experience to screen for the condition by obtaining simple two-dimension PLAX views without the need for comprehensive Doppler assessment, which can then be reserved for confirmatory assessment.

In the recent years a number of artificial intelligence applications have been described in the field of echocardiography,^31^ ranging from automated classification of echocardiographic views,^32^ video-based beat-to-beat assessment of left ventricular systolic dysfunction,^11^ detection of left ventricular hypertrophy and its various subtypes,^12^ diastolic dysfunction,^33^ to expert-level prenatal detection of complex congenital heart disease.^34^ Of note, machine learning methods further enable individuals without prior ultrasonography experience to obtain diagnostic TTE studies for limited diagnostic use.^35^ Despite this and even though the diagnosis and grading of AS remains dependent on echocardiography,^2,14^ most artificial intelligence solutions for timely AS screening have focused on alternative data types, such as audio files of cardiac auscultation,^36^ 12-lead electrocardiograms,^37–39^ cardio-mechanical signals using non-invasive wearable inertial sensors,^40^ as well as chest radiographs.^41^ For 12-lead electrocardiograms, AUC were consistently <0.90,^37–39^ whereas for alternative data types, analyses were limited to small datasets without external validation.^36,40^ Other studies have explored the value of structured data derived from comprehensive TTE studies in defining phenotypes with varying disease trajectories.^42^ More recently the focus has shifted to AI-assisted AS detection through automated TTE interpretation. In a recent study, investigators employed a form of self-supervised learning to automate the detection of AS, with their method however discarding temporal information by only including the first frame of each video loop, while also relying on the acquisition of images from several different views.^43^ The approach that relies on ultrasonography is also safer than the alternative screening strategies, such as those using chest computed tomography and aortic valve calcium scoring,^42,44^ which expose patients to radiation.

In this context, our work represents an advance both in the clinical and methodological space. First, we describe a method that can efficiently screen for a condition associated with significant morbidity and mortality,^2,7^ with increasing prevalence in the setting of an aging population.^45^ Our method has the potential to shift the initial burden away from trained echocardiographers and specialized core laboratories, as part of a more cost-effective screening and diagnostic cascade that can detect the condition at its earliest stages.^9,35^ In this regard, major strengths of our model include its reliance on a single echocardiographic view that can be obtained by individuals with limited experience and minimal training,^35^ and its ability to process temporal information through analysis of videos rather than isolated frames. The overarching goal is to develop screening tools that can be deployed in a cost-effective manner, gatekeeping access to comprehensive TTE assessment, which can be used as a confirmatory test to establish the suspected diagnosis.

Second, our work describes an end-to-end framework to boost artificial intelligence applications in echocardiography. We present an algorithm that automatically detects echocardiographic views, then performs self-supervised representation learning of PLAX videos with a multi-instance, contrastive learning approach. This novel approach further enables our algorithm to learn key representations of a patient’s cardiac phenotype that generalize and remain consistent across different clips and variations of the same echocardiographic views. By optimizing the detection of an echocardiographic fingerprint for each patient, this important pretraining step has the potential to boost AI-based echocardiographic assessment across a range of conditions. Furthermore, unlike previous approaches,^43^ our method benefits from multi-instance contrastive learning, which learns key representations using different videos from the same patient, a method that has been shown to improve predictive performance in the classification of dermatology images.^21^

Further to detecting severe AS, our algorithm learns features of aortic valvular pathology that generalize across different stages of the condition. Saliency maps demonstrate that the model focuses on the aortic valve with notable variation throughout the cardiac cycle, possibly learning features such as aortic valve calcification and restricted leaflet mobility.^14^ When restricting our analysis to patients without severe AS, the model’s predictions strongly correlated with Doppler-derived, quantitative features of stenosis severity. This is in accordance with the known natural history of AS, a progressive, degenerative condition, the hallmarks of which are aortic valve calcification, restricted mobility, functional stenosis and eventual ventricular decompensation.^2,7^ As such, our algorithm’s predictions also carry significant value as quantitative predictors of the stage of AV severity and could theoretically be used to monitor the rate of AS progression.

Limitations of our study include the lack of prospective validation of our findings. To this end, we are working on deploying this method in a prospective cohort of patients referred for routine TTE assessment to understand its real-world implications as a screening tool. Second, our model is limited to the use of PLAX views, which often represent the first step of TTE or POCUS protocols in cardiovascular assessment. Though there is no technical restriction to expanding these methods to alternative views, increasing the complexity of the screening protocol is likely to negatively impact its adoption in busy clinical settings. Finally, this study used data from formal TTEs, which generally produce higher-quality images than machines in POCUS settings. Though videos were downsampled for model development, further validation is needed to ensure robustness across acquisition technologies.

## CONCLUSION

In summary, we propose an efficient method to screen for severe AS using single-view (PLAX) TTE videos without the need for Doppler signals. More importantly, we describe an end-to-end approach for the deployment of artificial intelligence solutions in echocardiography, starting from automated view classification to self-supervised representation learning to accurate and explainable detection of severe AS. Our findings have significant implications for point-of-care ultrasound screening of AS as part of routine clinic visits and in limited resource settings and for individuals with minimal training.

## Data Availability

The data are not available for public sharing given the restrictions in our institutional review board approvals. A deidentified test set may be available to researchers under a data use agreement after the study has been published in a peer-reviewed journal.

## DATA AVAILABILITY

The data are not available for public sharing given the restrictions in our institutional review board approval. The deidentified test set may be available to researchers under a data use agreement after the study has been published in a peer-reviewed journal.

## CODE AVAILABILITY

The code repository for this work can be found at https://github.com/CarDS-Yale/echo-severe-AS.

## AUTHOR CONTRIBUTIONS

G.H. and E.K.O. performed the analyses, G.H., E.K.O. and R.K. drafted the manuscript, and all other authors provided critical revisions. A.C., N.Y., A.G., and D.O. facilitated cross-institution validation on data from Cedars-Sinai Hospital. R.K. supervised the study and is the guarantor.

