## Supplemental Materials for "Automated severe aortic stenosis detection on single-view echocardiography: A multi-center deep learning study"

**Supplemental methods:** pages 2-5

**Supplemental tables:** pages 6-7

**Supplemental figures:** pages 8

**Supplemental references:** pages 9-10

**Address for correspondence:**

Rohan Khera, MD, MS

195 Church St, 5^th^ Floor, New Haven, CT 06510

203-764-5885;; @rohan_khera

**Supplemental methods**

***Echocardiogram interpretation & labelling:*** The presence of AS severity was adjudicated based on the original echocardiographic report. Doppler assessment was interpreted based on the parameters recommended by the ASE, which included peak aortic valve (stenosis) jet velocity, mean transaortic/trans-valvular gradient, and mean valve area, as assess by continuity equation. According to the guidelines, cut-offs of >4 m/sec, >40 mm Hg and less than <1.0 cm^2^, respectively, were consistent with severe AS.^1^ The left ventricular ejection fraction (LVEF) was reported based on three-dimensional (3D) echocardiography, and in the absence of that, based on the Simpson’s biplane method. In the absence of these measurements, we reported the lower end of the visually estimated LVEF. Since severe AS detection was formulated as a binary classification task, all AS designations other than “severe AS” were binned into the “not severe AS” category.

***Data extraction and de-identification*:** In the New England cohort, after excluding studies that were not properly extracted from the database, 10,865 studies first underwent de-identification. After loading the pixel data for each video with the pydicom library (<https://pydicom.github.io/>), pixels in the periphery of each video frame were masked out to remove identifying information, and videos were converted to the Audio Video Interleave (AVI) format to enable fast loading for later preprocessing steps.

***View classification***: The resulting 447,653 videos from 9,710 studies then underwent view classification. Using the pretrained TTE view classifier from Zhang *et al.*,^2^ ten frames from each de-identified video were randomly selected, downsampled to 224 x 224 resolution, and fed through the pretrained VGG19 convolutional neural network. Video-level view predictions were then obtained by averaging each video’s 10 frame-wise view probabilities, and videos that were most confidently predicted as PLAX were kept for further preprocessing. While the pretrained view classifier was capable of discriminating variants of the standard PLAX view such as “PLAX”, “PLAX – remote,” “PLAX – zoom of left atrium,” and “PLAX – centered over left atrium,” we elected to only proceed with videos most confidently classified as “PLAX.”

***Initial video pre-processing:*** After view classification, the 30,136 videos from 9,173 studies were prepared for deep learning model development. Given differences in AS severity measures across different domains, we excluded echocardiograms with low-flow, low-gradient & paradoxical aortic stenosis leaving 29,978 PLAX videos from 9,122 studies. All videos underwent a more thorough cleaning and de-identification process that involved binarizing each video frame with a fixed threshold, then masking out all pixels outside the convex hull of the largest contour in order to remove all information outside the central image content. Finally, each video clip was spatially downsampled to 112 x 112 and saved to AVI format for fast loading during model training.

***Self-supervised contrastive learning:*** To learn transferable representations of PLAX echocardiogram videos for downstream severe AS identification, we performed self-supervised pretraining on all training set videos. This pretraining step critically enables the model to learn representations of echocardiograms that are robust to standard variations in video acquisition, thus better generalizing when later fine-tuned on a specific downstream task. We have previously demonstrated that a more appropriate initialization for data-efficient classification tasks could be achieved by “in-domain” pretraining on echocardiograms,^3^ as opposed to other standard approaches such as random initialization of weights and transfer learning.^4–6^ To this end, we have designed a novel self-supervised learning algorithm specifically catered to echocardiogram videos. More specifically, self-supervised representation learning was performed on the training set videos with a novel combination of (i) a multi-instance contrastive learning task and (ii) a frame re-ordering pretext task. In brief, we adopted the SimCLR framework^7^ for contrastive learning, which traditionally generates two “views” of an image by sending two copies of the input through a pipeline of random image augmentations, producing view $\tilde{x}_{i}$ and $\tilde{x}_{j}$. An encoder $f()$ is then used to learn representations of each view, $h_{i}=f(\tilde{x}_{i})$ and $h_{j}=f(\tilde{x}_{j})$, which are then projected to a lower dimensionality with a projector $g()$. The resulting learned embeddings of each view, $z_{i}=g(f\left( \tilde{x}_{i} \right))$and $z_{j}=g(f\left( \tilde{x}_{j} \right))$ are then “contrasted” via the temperature-normalized cross-entropy (NT-Xent) loss, which encourages the model to learn *similar* representations of views from the same original image (so-called “positive pairs”) and *dissimilar* representations of views from all other images (“negative pairs”) in a given minibatch. The model is then trained to learn *similar* representations of views from the same original image (so-called “positive pairs”) and *dissimilar* representations of views from all other images.

While SimCLR has proven very successful for 2D natural images as well as in medical applications such as radiography and dermatological images, there are several barriers to its successful adaptation to echocardiogram videos. First, SimCLR requires extremely heavy image augmentation for effective representation learning, which would destroy valuable signal encoded in the brittle, noisy ultrasound images produced by echocardiography. Second, SimCLR was designed for 2D images, which would completely ignore the temporal dimension of echocardiography. To address the first issue, we utilized “multi-instance” contrastive learning – borrowing language and key insights from Azizi *et al.*^8^ – whereby we form positive pairs between *different* videos from the *same* patient. This critically removes the need to synthetically create two different “views” of a patient by heavily augmenting their echo video, instead leveraging the fact that almost all studies contain multiple distinct PLAX videos of a patient.

To address the second issue, we additionally included a frame re-ordering “pretext” task to our self-supervised learning method, where we randomly permuted the frames of each input echo, then trained the model to predict the original order of frames. Similar to the approach of Jiao *et al.*,^9^ this frame re-ordering task is treated as a classification problem and was implemented with a simple fully-connected layer that minimizes the cross-entropy between the known and predicted original frame order; specifically, if an input echo clip has $K$ frames, then the $K!$ possible permutations of frames served as the targets for classification. Then the loss function of our self-supervised learning method is simply the sum of the contrastive NT-Xent objective and the pretext frame re-ordering cross-entropy objective. Self-supervised pretraining was performed on randomly sampled video clips of 4 consecutive frames from each of the training set echocardiogram videos for 300 epochs. The encoder $f()$ was a randomly initialized 3D-ResNet18,^10^ and the projector $g()$ projected each 512-dimensional learned representation down to a 128-dimensional representation with a hidden layer of 256 units followed by a ReLU activation, followed by an output layer with 128 units. The model was trained for 300 epochs on all unique pairs of *different* PLAX videos from the same patient with the Adam optimizer^11^ and a learning rate of 0.1, a batch size of 392 (196 per GPU), and NT-Xent temperature hyperparameter of 0.5. The following augmentations were applied to each frame in a temporally consistent manner (same transformations for each frame of a given video clip): random zero padding by up to 8 pixels in each spatial dimension, a random horizontal flip with probability 0.5, and a random rotation within -10 and 10 degrees with probability 0.5. After augmentation, each video clip was normalized so that the maximum pixel intensity was mapped to 1 and the minimum intensity to 0. The SSL model was trained on two NVIDIA RTX 3090 GPUs.

**Deep neural network training for severe AS prediction**

The same 3D-ResNet18 architecture was used to predict severe AS. Three different methods were used to initialize the parameters of this network: an SSL initialization, a Kinetics-400 initialization, and a random initialization. The SSL initialization directly used the learned weights of the encoder from the SSL pretraining step described in detail above. The Kinetics-400 initialization represents the “standard” transfer learning approach for 3D data, using the weights from a 3D-ResNet18 trained in a supervised fashion on the Kinetics-400 dataset, a large corpus of over 300,000 natural videos for human action classification; these weights are readily available through the torchvision API (<https://pytorch.org/vision/stable/index.html>) provided by PyTorch. The random initialization is the default when initializing a 3D-ResNet18 with PyTorch.^12^

All fine-tuning models were trained on randomly sampled video clips of 16 consecutive frames from training set echocardiograms, optionally padding with empty frames along the temporal axis if either the video is too short or the randomly chosen starting point of the clip is near the end of the video. The same augmentations were used as in self-supervised pretraining, and all video clips were min-max normalized; when fine-tuning from a Kinetics-400 initialization, video clips were further standardized using the channel-wise means and standard deviations from the Kinetics-400 training dataset, a standard preprocessing step when performing transfer learning. All models were trained for a maximum of 30 epochs with early stopping – specifically, if the validation AUROC did not improve for 5 consecutive epochs, training was terminated and the weights from the epoch with maximum validation AUROC were used for final evaluation. Severe AS models were trained on a single NVIDIA RTX 3090 GPU with the Adam optimizer, a learning rate of $1\times{10}^{-4}$ (except the SSL-pretrained model, which used a learning rate of 0.1), and a batch size of 88 in order to maximize GPU utilization. Since this problem was framed as a binary classification task, these models minimized a sigmoid cross-entropy loss. We additionally used class weights computed with the method provided by scikit-learn^13^ to accommodate class imbalance in addition to label smoothing^14,15^ with $\alpha$=0.1, a method to improve model calibration and generalization. Learning curves depicting loss throughout training were graphically visualized.

**Ensembling:** We formed an ensemble of three models trained to detect severe AS, with diversity injected by the dramatically different methods used to initialize each model’s weights before training. Ensembling is known to improve predictive performance by aggregating the outputs of multiple independently trained models.^16^ Moreover, statistical^17–19^ and deep learning^20–22^ studies have shown that ensembles of *diverse* constituent models are most effective.

More specifically, since the fine-tuned severe AS models are trained on 16-frame video clips, yet AS labels describe each study, we first aggregated clip-level predictions into study-level predictions for performance evaluation. When performing inference on an echo video, four evenly spaced clips (potentially with overlapping frames) of 16 consecutive frames were extracted and fed into the trained AS model. These clip-level predictions were then averaged to obtain a video-level prediction of severe AS. After repeating this process for all videos, the severe AS probabilities for videos in each study were averaged to obtain study-level AS predictions that could be used to compute evaluation metrics. The final ensemble model is then formed by averaging the output probabilities of the SSL-pretrained model, the Kinetics-400-pretrained model, and the randomly initialized model after fine-tuning each ensemble member to classify severe AS. Since no quality control is applied when selecting PLAX videos for this work, averaging results over multiple videos in the same study has a stabilizing effect that boosts predictive performance.

**Model explainability:** Saliency maps were generated by leveraging the Grad-CAM method^23^ for obtaining visual explanations from deep neural networks. Specifically, heatmaps were generated by applying Grad-CAM to a clip of the first 32 frames of an echo, using the last convolution block of the 3D ResNet18 to generate a 7 x 7 x 4 (height x width x time) heatmap displaying roughly where the model is attending to over the spatial and temporal dimensions. The Grad-CAM output was interpolated to the original input dimension of 112 x 112 x 32 with the scipy “zoom” function; this process produces a frame-by-frame “visual explanation” of *where* the model is focusing frame by frame in order to make its prediction. However, to generate a single 2D heatmap for a given echo clip, the pixelwise maximum along the temporal axis was taken to capture the most salient regions for severe AS predictions across all timepoints.

**Supplemental tables**

**Table S1 | External performance of an automated algorithm for detection of aortic stenosis.**

|  | **SSL** | **Kinetics-400** | **Random** | **Ensemble** |
| --- | --- | --- | --- | --- |
| **External testing set from New England/YNHHS (2016-2020)** | | | | |
| **AUROC** | 0.911 (0.891, 0.930) | 0.908 (0.889, 0.927) | 0.903 (0.882, 0.923) | **0.915 (0.896, 0.933)** |
| **AUPR** | 0.652 (0.580, 0.723) | 0.621 (0.548, 0.697) | 0.615 (0.542, 0.687) | **0.654 (0.585, 0.721)** |
| **F1 Score** | 0.647 (0.608, 0.706) | 0.640 (0.603, 0.697) | 0.636 (0.594, 0.696) | **0.656 (0.620, 0.713)** |
| **PPV** | 0.595 (0.494, 0.733) | 0.569 (0.479, 0.635) | 0.628 (0.482, 0.697) | **0.582 (0.498, 0.688)** |
| **Sensitivity** | 0.709 (0.598, 0.894) | 0.730 (0.690, 0.901) | 0.645 (0.605, 0.887) | **0.752 (0.652, 0.910)** |
| **Specificity at 90% Sensitivity** | 0.806 (0.709, 0.836) | 0.804 (0.700, 0.843) | 0.783 (0.720, 0.822) | **0.824 (0.725, 0.857)** |
| **PPV at 90% Sensitivity** | 0.476 (0.369, 0.531) | 0.472 (0.369, 0.538) | 0.447 (0.378, 0.510) | **0.500 (0.379, 0.564)** |
| **Temporally distinct testing set from YNHHS (2021)** | | | | |
| **AUROC** | 0.969 (0.951, 0.984) | 0.979 (0.971, 0.987) | 0.969 (0.948, 0.985) | **0.978 (0.966, 0.988)** |
| **AUPR** | 0.270 (0.144, 0.470) | 0.305 (0.160, 0.484) | 0.372 (0.193, 0.555) | **0.414 (0.231, 0.594)** |
| **F1 Score** | 0.400 (0.276, 0.588) | 0.417 (0.293, 0.566) | 0.429 (0.294, 0.600) | **0.431 (0.323, 0.619)** |
| **PPV** | 0.600 (0.205, 0.833) | 0.357 (0.214, 0.625) | 0.409 (0.237, 1.000) | **0.355 (0.263, 0.875)** |
| **Sensitivity** | 0.300 (0.238, 0.700) | 0.500 (0.312, 0.762) | 0.450 (0.250, 0.682) | **0.550 (0.286, 0.720)** |
| **Specificity at 90% Sensitivity** | 0.943 (0.800, 0.962) | 0.957 (0.943, 0.965) | 0.945 (0.787, 0.963) | **0.954 (0.880, 0.971)** |
| **PPV at 90% Sensitivity** | 0.135 (0.041, 0.207) | 0.171 (0.116, 0.237) | 0.140 (0.039, 0.212) | **0.162 (0.067, 0.248)** |
| **External testing set from Cedars-Sinai (2018-2019)** | | | | |
| **AUROC** | 0.965 (0.962, 0.969) | 0.967 (0.963, 0.970) | 0.953 (0.948, 0.957) | **0.972 (0.969, 0.975)** |
| **AUPR** | 0.922 (0.912, 0.932) | 0.917 (0.906, 0.928) | 0.893 (0.881, 0.904) | **0.932 (0.921, 0.941)** |
| **F1 Score** | 0.865 (0.856, 0.876) | 0.865 (0.857, 0.877) | 0.831 (0.821, 0.843) | **0.880 (0.871, 0.890)** |
| **PPV** | 0.841 (0.807, 0.866) | 0.846 (0.805, 0.866) | 0.796 (0.763, 0.813) | **0.851 (0.832, 0.865)** |
| **Sensitivity** | 0.889 (0.865, 0.929) | 0.886 (0.868, 0.934) | 0.869 (0.857, 0.908) | **0.911 (0.900, 0.930)** |
| **Specificity at 90% Sensitivity** | 0.917 (0.906, 0.929) | 0.918 (0.908, 0.928) | 0.881 (0.865, 0.891) | **0.932 (0.924, 0.940)** |
| **PPV at 90% Sensitivity** | 0.070 (0.067, 0.073) | 0.074 (0.071, 0.076) | 0.065 (0.063, 0.068) | **0.075 (0.073, 0.077)** |
| Results come from a 3D-ResNet18 when initialized with the proposed self-supervised learning (SSL) initialization, a standard transfer learning approach (Kinetics-400), and a random weight initialization. “Ensemble” denotes an ensemble of the three individual models described above. Values in parentheses represent 95% confidence intervals determined by bootstrapping. AUROC = area under the receiver operating characteristic curve; AUPR = area under the precision-recall curve; PPV = positive predictive value; YNHHS = Yale-New Haven Health System. | | | | |

**Table S2 | External performance of an automated algorithm for detection of aortic stenosis on individual PLAX videos.**

|  | **SSL** | **Kinetics-400** | **Random** | **Ensemble** |
| --- | --- | --- | --- | --- |
| **External testing set from New England/YNHHS (2016-2020)** | | | | |
| **AUROC** | 0.877 (0.863, 0.891) | 0.884 (0.871, 0.897) | 0.867 (0.853, 0.881) | **0.887 (0.874, 0.900)** |
| **AUPR** | 0.575 (0.532, 0.619) | 0.583 (0.540, 0.627) | 0.524 (0.480, 0.567) | **0.594 (0.552, 0.635)** |
| **F1 Score** | 0.570 (0.544, 0.603) | 0.576 (0.546, 0.611) | 0.545 (0.520, 0.580) | **0.579 (0.555, 0.614)** |
| **PPV** | 0.476 (0.439, 0.555) | 0.569 (0.491, 0.607) | 0.444 (0.416, 0.511) | **0.473 (0.444, 0.571)** |
| **Sensitivity** | 0.709 (0.602, 0.782) | 0.583 (0.555, 0.671) | 0.706 (0.599, 0.756) | **0.748 (0.608, 0.800)** |
| **Specificity at 90% Sensitivity** | 0.666 (0.624, 0.698) | 0.719 (0.676, 0.758) | 0.671 (0.637, 0.702) | **0.698 (0.662, 0.745)** |
| **PPV at 90% Sensitivity** | 0.319 (0.287, 0.348) | 0.357 (0.320, 0.397) | 0.322 (0.294, 0.352) | **0.342 (0.311, 0.387)** |
| **Temporally distinct testing set from YNHHS (2021)** | | | | |
| **AUROC** | 0.951 (0.937, 0.964) | 0.959 (0.948, 0.969) | 0.942 (0.921, 0.960) | **0.961 (0.949, 0.972)** |
| **AUPR** | 0.236 (0.153, 0.346) | 0.251 (0.162, 0.352) | 0.264 (0.169, 0.368) | **0.353 (0.246, 0.464)** |
| **F1 Score** | 0.360 (0.278, 0.466) | 0.347 (0.272, 0.451) | 0.340 (0.277, 0.455) | **0.389 (0.312, 0.500)** |
| **PPV** | 0.400 (0.250, 0.613) | 0.318 (0.221, 0.500) | 0.410 (0.214, 0.538) | **0.396 (0.284, 0.786)** |
| **Sensitivity** | 0.327 (0.241, 0.482) | 0.382 (0.268, 0.522) | 0.291 (0.262, 0.569) | **0.382 (0.250, 0.516)** |
| **Specificity at 90% Sensitivity** | 0.848 (0.831, 0.903) | 0.890 (0.845, 0.920) | 0.840 (0.733, 0.895) | **0.890 (0.840, 0.916)** |
| **PPV at 90% Sensitivity** | 0.050 (0.040, 0.072) | 0.066 (0.045, 0.094) | 0.046 (0.027, 0.068) | **0.065 (0.045, 0.089)** |
| **External testing set from Cedars-Sinai (2018-2019)** | | | | |
| **AUROC** | 0.948 (0.945, 0.951) | 0.953 (0.950, 0.956) | 0.926 (0.922, 0.929) | **0.958 (0.956, 0.961)** |
| **AUPR** | 0.891 (0.884, 0.899) | 0.895 (0.887, 0.902) | 0.853 (0.844, 0.861) | **0.908 (0.901, 0.915)** |
| **F1 Score** | 0.817 (0.811, 0.825) | 0.835 (0.829, 0.843) | 0.783 (0.777, 0.792) | **0.842 (0.835, 0.849)** |
| **PPV** | 0.780 (0.765, 0.834) | 0.797 (0.791, 0.835) | 0.758 (0.724, 0.768) | **0.812 (0.801, 0.834)** |
| **Sensitivity** | 0.859 (0.803, 0.878) | 0.876 (0.837, 0.883) | 0.811 (0.803, 0.852) | **0.874 (0.848, 0.885)** |
| **Specificity at 90% Sensitivity** | 0.850 (0.839, 0.861) | 0.876 (0.866, 0.884) | 0.789 (0.776, 0.801) | **0.883 (0.874, 0.892)** |
| **PPV at 90% Sensitivity** | 0.063 (0.061, 0.064) | 0.066 (0.065, 0.068) | 0.055 (0.054, 0.057) | **0.068 (0.066, 0.069)** |
| Results come from a 3D-ResNet18 when initialized with the proposed self-supervised learning (SSL) initialization, a standard transfer learning approach (Kinetics-400), and a random weight initialization. “Ensemble” denotes an ensemble of the three individual models described above. Values in parentheses represent 95% confidence intervals determined by bootstrapping. AUROC = area under the receiver operating characteristic curve; AUPR = area under the precision-recall curve; PPV = positive predictive value; YNHHS = Yale-New Haven Health System. | | | | |

**Supplemental figures**

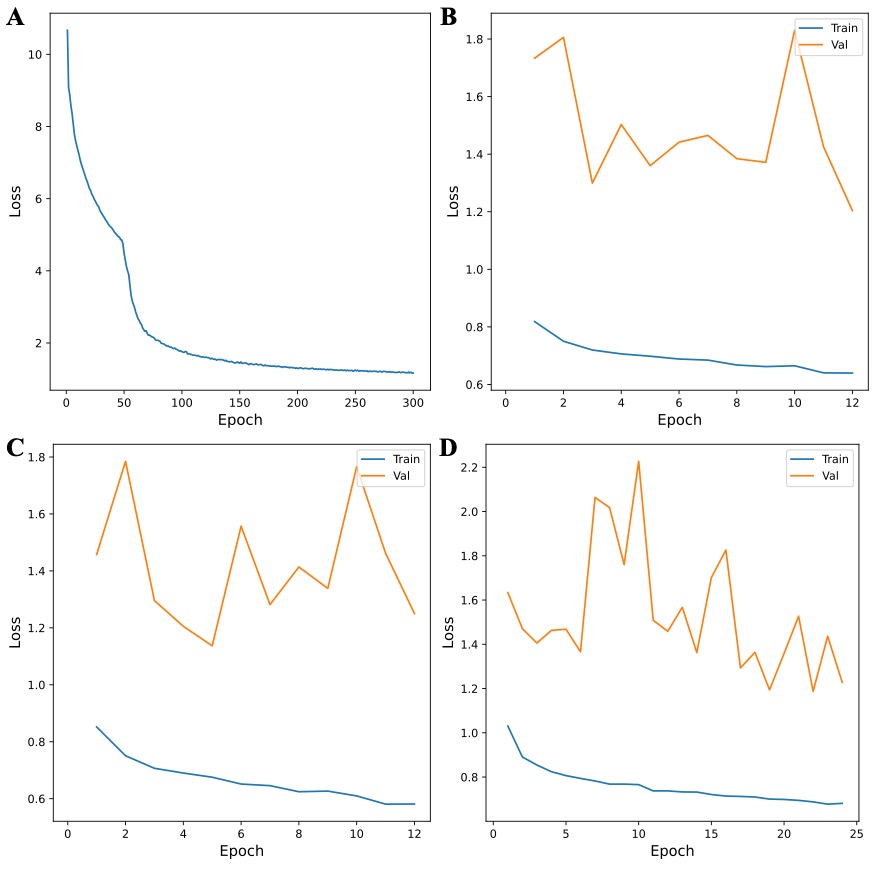

**Figure S1 | Learning curves during model training.** Training curves depicting model loss throughout self-supervised learning (SSL) (a), fine-tuning from a random initialization (b), fine-tuning from a Kinetics-400 initialization (c), and fine-tuning from our SSL initialization (d).

3.	Holste G, Oikonomou EK, Mortazavi B, Wang Z, Khera R. Self-supervised learning of echocardiogram videos enables data-efficient clinical diagnosis. *arXiv [cs.CV]*.

4.	Rajpurkar P, Irvin J, Zhu K, Yang B, Mehta H, Duan T, Ding D, Bagul A, Langlotz C, Shpanskaya K, Lungren MP, Ng AY. CheXNet: Radiologist-Level Pneumonia Detection on Chest X-Rays with Deep Learning. *arXiv [cs.CV]*.

5.	Gulshan V, Peng L, Coram M, Stumpe MC, Wu D, Narayanaswamy A, Venugopalan S, Widner K, Madams T, Cuadros J, Kim R, Raman R, Nelson PC, Mega JL, Webster DR. Development and Validation of a Deep Learning Algorithm for Detection of Diabetic Retinopathy in Retinal Fundus Photographs. *JAMA* 2016;**316**:2402–2410.

6.	Ouyang D, He B, Ghorbani A, Yuan N, Ebinger J, Langlotz CP, Heidenreich PA, Harrington RA, Liang DH, Ashley EA, Zou JY. Video-based AI for beat-to-beat assessment of cardiac function. *Nature* 2020;**580**:252–256.

7.	Chen T, Kornblith S, Norouzi M, Hinton G. A Simple Framework for Contrastive Learning of Visual Representations. Iii HD, Singh A, eds. *Proceedings of the 37th International Conference on Machine Learning*. PMLR; 13--18 Jul 2020. p1597–1607.

8.	Azizi S, Mustafa B, Ryan F, Beaver Z, Freyberg J, Deaton J, Loh A, Karthikesalingam A, Kornblith S, Chen T, Natarajan V, Norouzi M. Big self-supervised models advance medical image classification. *2021 IEEE/CVF International Conference on Computer Vision (ICCV)*. IEEE; 2021.

9.	Jiao J, Droste R, Drukker L, Papageorghiou AT, Noble JA. Self-Supervised Representation Learning for Ultrasound Video. *Proc IEEE Int Symp Biomed Imaging* 2020;**2020**:1847–1850.

10.	Tran, Wang, Torresani, Ray. A closer look at spatiotemporal convolutions for action recognition. *Proc Estonian Acad Sci Biol Ecol* 2018.

11.	Kingma DP, Ba J. Adam: A Method for Stochastic Optimization. *arXiv [cs.LG]*.

12.	Paszke, Gross, Massa, Lerer. Pytorch: An imperative style, high-performance deep learning library. *Adv Neural Inf Process Syst* 2019.

13.	Pedregosa, Varoquaux, Gramfort. Scikit-learn: Machine learning in Python. *the Journal of machine* 2011.

14.	Müller, Kornblith, Hinton. When does label smoothing help? *Adv Neural Inf Process Syst* 2019.

15.	Szegedy, Vanhoucke, Ioffe. Rethinking the inception architecture for computer vision. *Proc Estonian Acad Sci Biol Ecol* 2016.

16.	Dietterich TG. Ensemble Methods in Machine Learning. *Multiple Classifier Systems*. Springer Berlin Heidelberg; 2000. p1–15.

17.	Dietterich TG. An Experimental Comparison of Three Methods for Constructing Ensembles of Decision Trees: Bagging, Boosting, and Randomization. *Mach Learn* 2000;**40**:139–157.

18.	Opitz, Shavlik. Generating accurate and diverse members of a neural-network ensemble. *Adv Neural Inf Process Syst* 1995.

19.	Brown, Wyatt, Tino, Bengio. Managing diversity in regression ensembles. *J Mach Learn Res* 2005.

20.	Wasay, Hentschel, Liao. Mothernets: Rapid deep ensemble learning. *of Machine Learning …* 2020.

21.	Lakshminarayanan, Pritzel. Simple and scalable predictive uncertainty estimation using deep ensembles. *Adv Neural Inf Process Syst* 2017.

22.	Qummar S, Khan FG, Shah S, Khan A, Shamshirband S, Rehman ZU, Ahmed Khan I, Jadoon W. A Deep Learning Ensemble Approach for Diabetic Retinopathy Detection. *IEEE Access* 2019;**7**:150530–150539.

23.	Selvaraju RR, Cogswell M, Das A, Vedantam R, Parikh D, Batra D. Grad-CAM: Visual Explanations from Deep Networks via Gradient-based Localization. *arXiv [cs.CV]*.
